# SARS-CoV-2 viral load monitoring by extraction-free testing of saliva

**DOI:** 10.1101/2021.08.02.21261502

**Authors:** Yue Qiu, Ling Lu, Dexiang Gao, Patrick McGrath, Chann Han, Igor Kogut, Bob Blomquist, Xin Yao, Jose P. Zevallos, Brian L. Harry, Shi-Long Lu

## Abstract

Real-time quantitative reverse transcriptase polymerase chain reaction (RT-qPCR) remains the foundation of SARS-CoV-2 testing due to its accessibility, scalability, and superior assay performance. Variability in specimens and methods prevent standardization of RT-qPCR assays and reliable quantitative reporting to assess viral load. We developed an extraction-free RT-qPCR assay for detection of SARS-CoV-2 in saliva and monitored viral load until convalescence in COVID-19 patients. Comparison of 231 matched anterior nares swab and saliva specimens demonstrated that extraction-free testing of saliva has equivalent analytical and clinical assay performance compared to testing of RNA extracts from either anterior nares or saliva specimens. Analysis of specimen pairs revealed higher viral loads in the nasal cavity compared to the oral cavity, although this difference did not impact clinical sensitivity for COVID-19. Extraction-free testing of a combination specimen consisting of both nasal swab and saliva is also demonstrated. Assessment of viral load by RT-qPCR and parallel digital droplet PCR (ddPCR) revealed that cycle threshold (Ct) values less than approximately 30 correlated well with viral load, whereas Ct values greater than 30 correspond to low viral loads <10 copies/µL. Therefore, extraction-free saliva testing maximizes testing efficiency without compromising assay performance and approximates viral loads >10 copies/µL. This technology can facilitate high-throughput laboratory testing for SARS-CoV-2, monitor viral load in individual patients, and assess efficacy of therapies for COVID-19.

## Introduction

The COVID-19 pandemic has necessitated rapid scaling of laboratory and testing resources globally. Real-time quantitative reverse transcriptase polymerase chain reaction (RT-qPCR) is an accessible and scalable technology with excellent performance to detect SARS-CoV-2. RT-qPCR remains the foundation of testing for COVID-19.

Clinical RT-qPCR assays for SARS-CoV-2 depend on many variables. Specimen types, collection devices, storage and transport conditions, time from collection, nucleic acid targets, primers and probes, extraction method, amplification method, instrumentation, and other factors contribute to a high degree of inter-assay variability. Proficiency testing surveys using single-batch material with low viral load revealed that the median Ct value spans 14 cycles across laboratories and precision spans 3 cycles on a single instrument (*1*), which are unacceptable for viral load quantitation. Accordingly, the FDA has issued Emergency Use Authorization (EUA) only for qualitative interpretation of nucleic acid amplification (*2*). Yet some studies suggest that viral load is important for transmissibility, risk stratification, and prognosis in select COVID-19 patient populations (*3-7*). Therefore, precise viral load quantitation is a challenge and an opportunity to enhance the utility of SARS-CoV-2 RT-qPCR methods.

RNA extraction involves cell lysis using inhibitors of RNase activity, separation of RNA from other macromolecules, and concentration of RNA. During the COVID-19 pandemic, this process has been hindered by strained supply chains and has compounded inefficiency in high-throughput testing. Thus, RNA extraction by non-commercial methods and extraction-free testing have been explored as innovative approaches to address a major clinical need (*8-13*). Given that RNA extraction increases the operational and financial cost of the testing, it limits scalability of testing. Extraction-free testing is an attractive alternative in both resource-limited and high-throughput laboratories alike.

Nasopharyngeal swab (NP) swab specimens initially were the gold standard for the detection of SARS-CoV-2 early in the COVID-19 pandemic. Alternative specimen types from the upper respiratory tract (e.g., mid-turbinate, anterior nares, oropharyngeal, saliva) and lower respiratory tract (e.g., sputum, tracheal aspirate, bronchioalveolar lavage) have been validated against paired NP swabs with a high degree of concordance (*8, 14*). Discordance between paired specimens from the same patient is often associated with low viral load (*14*). Anterior nares swabs and saliva specimens are particularly attractive because they are less invasive and easier to collect compared to. Therefore, nasal swab and saliva have thus emerged as important specimen types for the diagnosis of COVID-19.

We developed a SARS-CoV-2 RT-qPCR assay and performed viral load monitoring of non-hospitalized COVID-19 patients until convalescence. Comparison of 231 matched anterior nares swab (hereafter nasal swab) and saliva specimens collected longitudinally demonstrated a high degree of qualitative agreement with or without RNA extraction. Viral load by RT-qPCR was benchmarked against digital droplet PCR (ddPCR) to confirm the quantitative value of RT-qPCR for viral load monitoring.

## Results

### Analytical validation of extraction-free testing

We modified the 2019-nCoV CDC real-time RT-qPCR assay, targeting the nucleocapsid gene of SARS-CoV-2 (N1 and N2) and the human ribonuclease P (RNP) gene, for extraction-free testing of saliva (*15*). Assay performance was compared using RNA extract from nasal swab, RNA extract from saliva, unprocessed saliva, and unprocessed saliva combined with nasal swab at the time of collection – designated as SwabCLEAR™, SalivaCLEAR™, SalivaFAST™, and Spit-N-Swab™. We focused on development of SalivaFAST for extraction-free saliva testing. We focused on validation of SalivaFAST.

Analytical validation was performed by spiking COVID-19-negative nasal swab and saliva specimens with varying concentrations of SARS-CoV-2 positive control material (2-100 genome equivalents per microliter, GE/µL). Assay precision at these low viral loads corresponded to N1 Ct values >30 with coefficients of variation spanning 1.02-4.26% for SwabCLEAR and 0.95-4.33% for SalivaFAST and, respectively (Fig. 1A). Compared to SwabCLEAR, SalivaFAST generally had higher Ct values at low viral loads, suggesting signal enhancement from RNA extraction of nasal swabs or inhibition by saliva matrix. As expected RNP Ct values, which are based on an intrinsic host target, were more variable but unaffected by SARS-CoV-2 (Fig. 1B). The SalivaFAST limit of detection (LoD) was determined to be 4 GE/µL (Fig. 1C). Analytical specificity was established previously (*15, 16*). These data suggest that extraction-free saliva testing has acceptable analytical performance compared to RNA extract from nasal swab.

**Figure 1.**
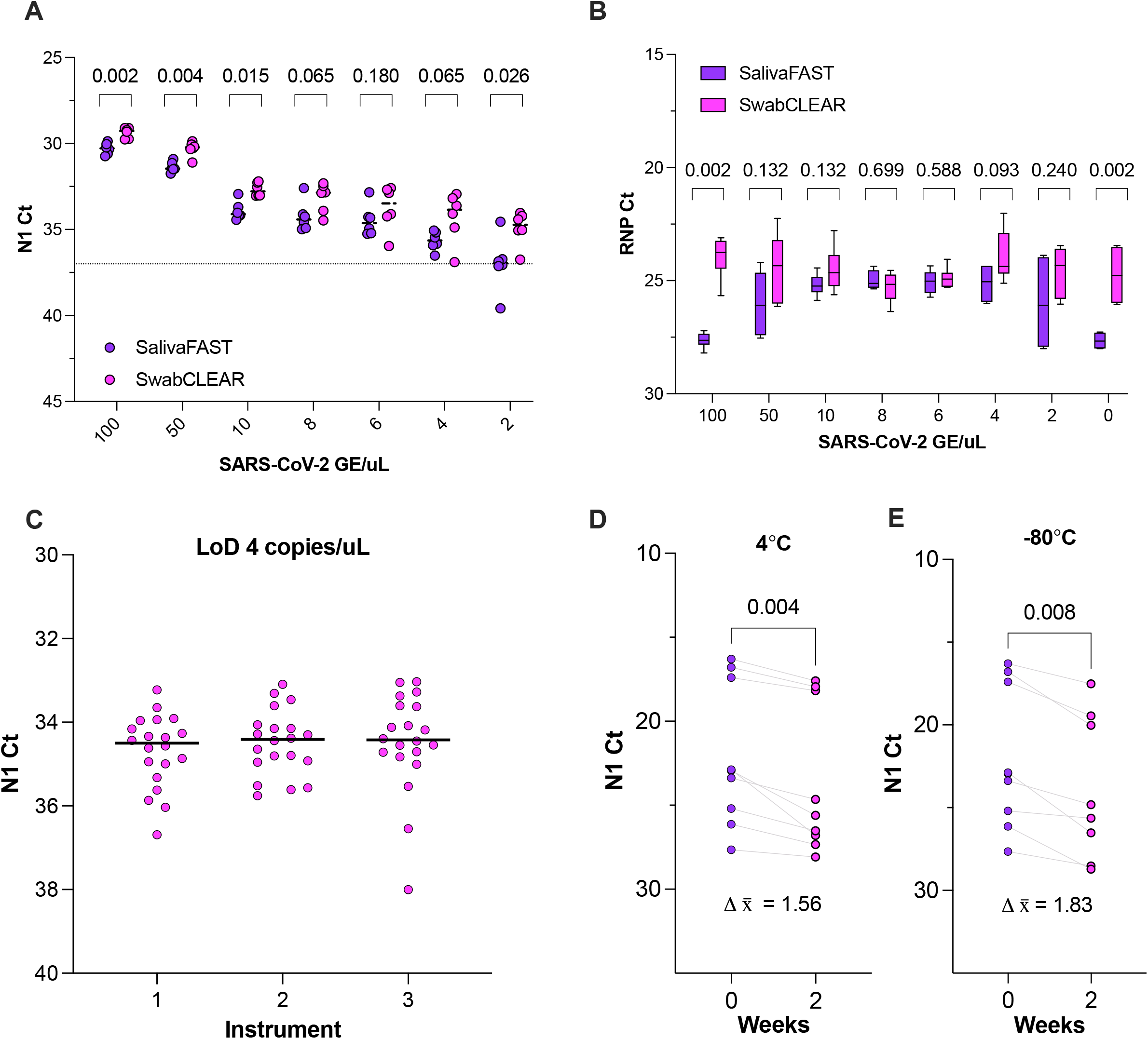
Analytical validation of extraction-free saliva testing. RT-qPCR using negative saliva (SalivaFAST) and nasal swabs (SwabCLEAR) spiked with SARS-CoV-2 control material demonstrate similar performance for N1 (A) and RNP (B). The limit of detection (LoD) of SalivaFAST was confirmed by analysis of 20 samples at 4 GE/µL on 3 separate thermocyclers (C). SalivaFAST performed at 0 and 2 weeks demonstrated a slight decreased viral load at both 4°C (D) and -80°C (E) although this did not impact clinical sensitivity.

The pre-analytical phase from specimen collection to laboratory testing is complicated by variables related to specimens, transport, and environmental conditions. We assessed the stability of samples tested by SalivaFAST after 2 weeks. The amount of detectable SARS-CoV-2 RNA diminished, with a mean N1 Ct increase of 1.56 and 1.83 for specimens stored at 4°C and -80°C, respectively (Fig. 1D-E) (n=8, *p* = 0.004 and *p* = 0.008).

### Clinical validation of RNA extracts from nasal swab and saliva specimens

The performance of the 2019-nCoV CDC real-time RT-qPCR assay has been extensively studied and validated using RNA extracts from nasal swabs. To begin clinical validation of SalivaFAST, we collected matched nasal swab and saliva specimens prospectively from 137 patients of which 19 (13.9%) were diagnosed with COVID-19. COVID-19 patients underwent longitudinal testing until convalescence (Fig. S1), yielding 231 matched sample sets. All specimen types demonstrated viral loads throughout the detectable range, including an abundance of low viral load specimens with N1 Ct values >30 (Fig. 2A-B). The RNP host gene signal was highly consistent across all specimen types regardless of COVID-19 status (Fig. 2B).

**Figure 2.**
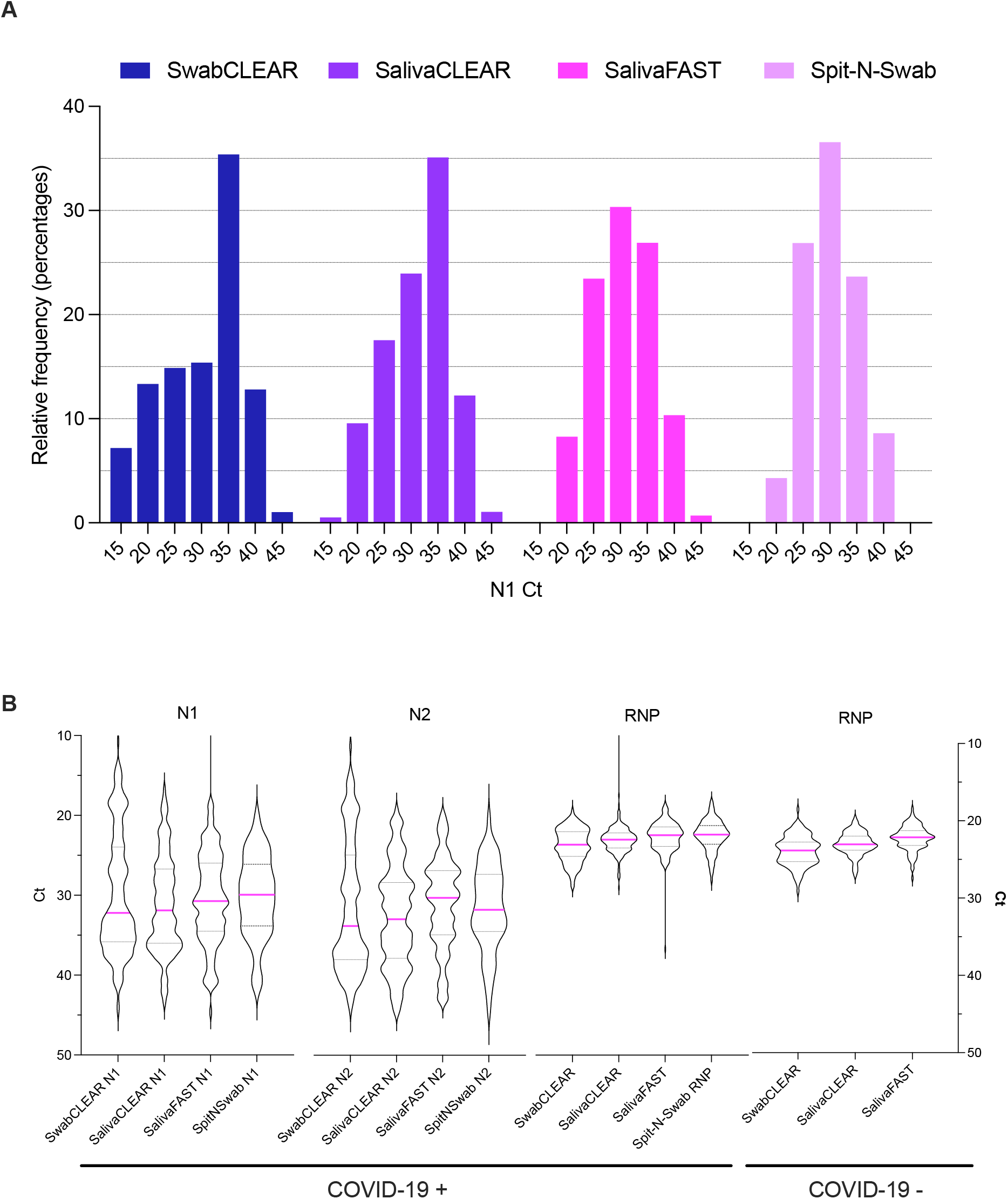
Distribution of Ct values from clinical samples. Matched RNA extracts from nasal swab (SwabCLEAR), RNA extracts from saliva (SalivaCLEAR), extraction-free saliva (SalivaFAST), and extraction-free saliva with a combined nasal swab specimen (Spit-N-Swab) demonstrate viral loads across a broad range according to the N1 Ct value, with an abundance of low viral load samples (A). Violin plots demonstrate the distribution of Ct values for the N1, N2, and RNP targets from all 3 assays (B).

To understand the impact of specimen type on SARS-CoV-2 RT-qPCR testing, N1 Ct values were compared between matched samples tested by SwabCLEAR and SalivaCLEAR, both of which depend on RNA extraction. Comparison of initial diagnostic samples revealed a mean Ct value increase of 5.87 by SalivaCLEAR (n=19 sample pairs, *p* < 0.001). By day 5 this difference decreased to 1.74 (n=16 sample pairs, *p* = 0.034) (Fig. 3C-D). Throughout the disease course from diagnosis to convalescence, SwabCLEAR revealed a slightly higher viral load compared to SalivaCLEAR (29.71 ± 0.30 versus 31.23 ± 0.24, n = 229 sample pairs, *p* < 0.001, Fig. 3E). Therefore, viral load by RT-qPCR is higher in the nasal cavity compared to the oral cavity and this difference diminishes over the course of infection.

**Figure 3.**
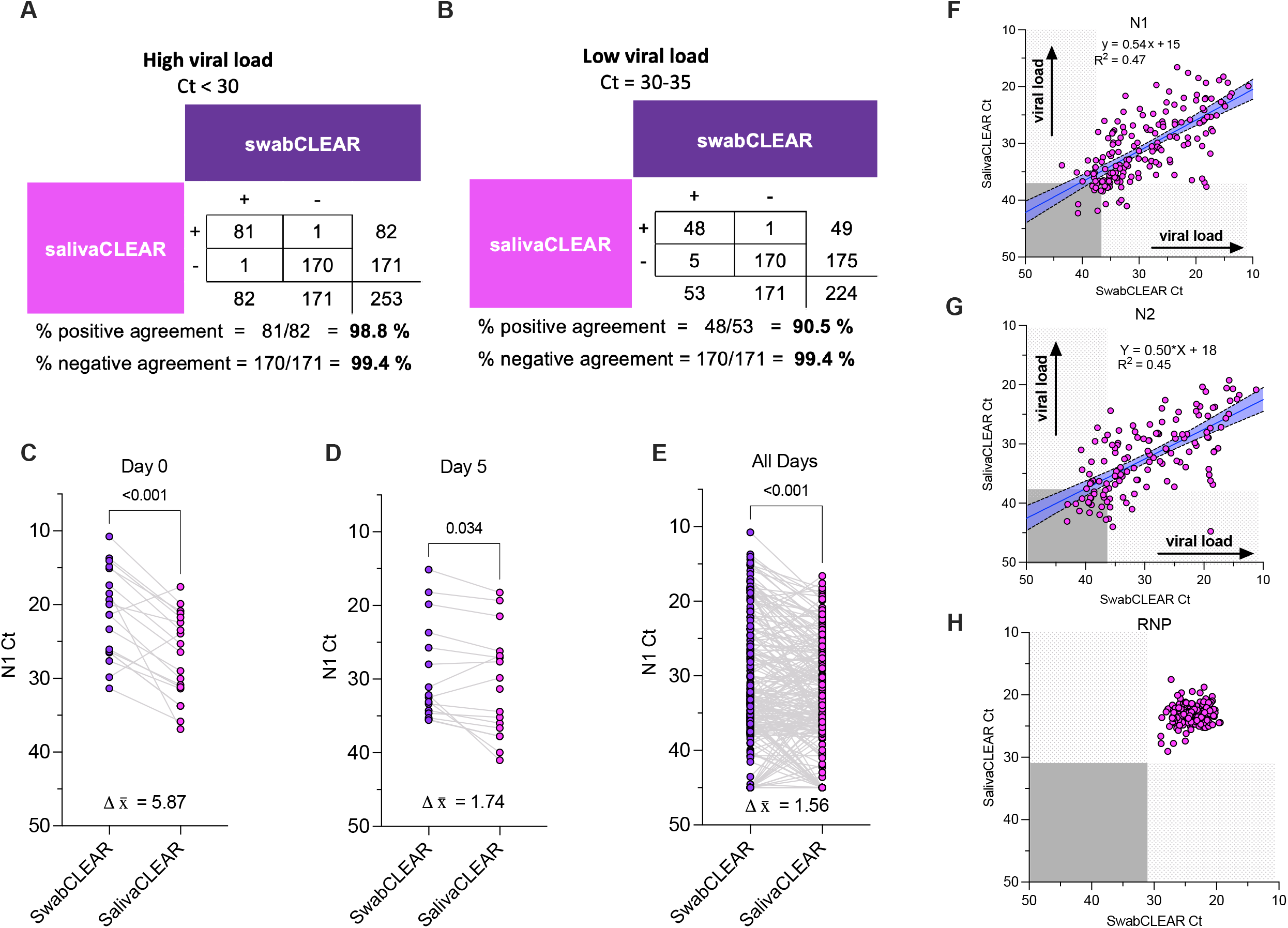
Impact of biospecimen type on RT-qPCR-based detection of SARS-CoV-2. Positive and negative agreement between matched nasal swab (SwabCLEAR) and saliva (SalivaCLEAR) following RNA extraction was assessed for high viral load nasal swab samples with a Ct<30 (A) and low viral load nasal swab samples Ct = 30-35 (B). Comparison of N1 Ct values from matched SwabCLEAR and SalivaCLEAR samples demonstrated lower viral load in saliva at day 0 (C), day 5 (D), and all timepoints during longitudinal monitoring (E). There was moderate correlation between N1 Ct values (F) and N2 Ct values (G) for SwabCLEAR and SalivaCLEAR. RNP values are consistent for SwabCLEAR and SalivaCLEAR (H).

A difference in viral load between specimen types has the potential to impact clinical sensitivity. Positive and negative agreement between SwabCLEAR and SalivaCLEAR specimens were assessed according to viral load. We defined high viral load as Ct <30, corresponding to >100 GE/µL (Fig. 1A). High viral load was associated with 98.8% positive agreement. Low viral load specimens demonstrated 88.7% positive agreement (Fig. 3A-B). Negative agreement in matched specimens from patients without COVID-19 was >99% at both levels. Moderate correlation was observed between N1 and N2 in each assay (*R*^*2*^ =0.45-0.47, Fig. 3F-G) whereas the RNP Ct values were similar and consistent between specimens (Fig. 3H, *p* = 0.839). These data suggest that there is excellent diagnostic agreement between RNA extracts from paired nasal swab and saliva specimens for diagnosis of COVID-19 despite higher viral load of SARS-CoV-2 in the nasal cavity.

### Clinical validation of extraction-free saliva testing

Supply chain disturbances during the pandemic have limited the availability of critical laboratory supplies, such as viral transport media and collection devices. RNA extraction-free RT-qPCR testing has emerged as an attractive alternative, including direct testing of saliva (*8, 10*). Since saliva collected in commercial device buffers could not be amplified without prior RNA extraction (data not shown), an additional saliva sample was collected in an inert plastic tube without buffer for SalivaFAST testing. RNA extraction of saliva resulted in a significant but negligible increase in viral load, with a mean increased N1 Ct value of 0.07 for SalivaCLEAR compared to SalivaFAST (Fig. 4C, n = 202 sample pairs, *p* = 0.046). Positive agreement between SalivaCLEAR and SalivaFAST varied according to viral load. Saliva specimens with high viral load (N1 Ct value ≤ 30) demonstrated 100.0% percent positive agreement between SalivaCLEAR and SalivaFAST (Fig. 4A, n=65 sample pairs). Saliva specimens with low viral load (N1 Ct value 30-35) demonstrated 81.8% positive agreement (Fig. 4B, n=44). Additionally, in some specimens with low viral load, SalivaCLEAR and SalivaFAST each detected SARS-CoV-2 not detected by the alternative method (Fig. 4B), suggesting that RNA extraction represents a trade-off between RNA concentration and RNA yield. Comparison of N1 and N2 between assays again demonstrated moderate correlation (Fig. 3D-E, *R*^*2*^ =0.58 for N1, *R*^*2*^ =0.40 for N2). These data demonstrate that RNA extraction is not necessary for reliable COVID-19 diagnosis using saliva and that extraction-free testing does not compromise clinical sensitivity.

**Figure 4.**
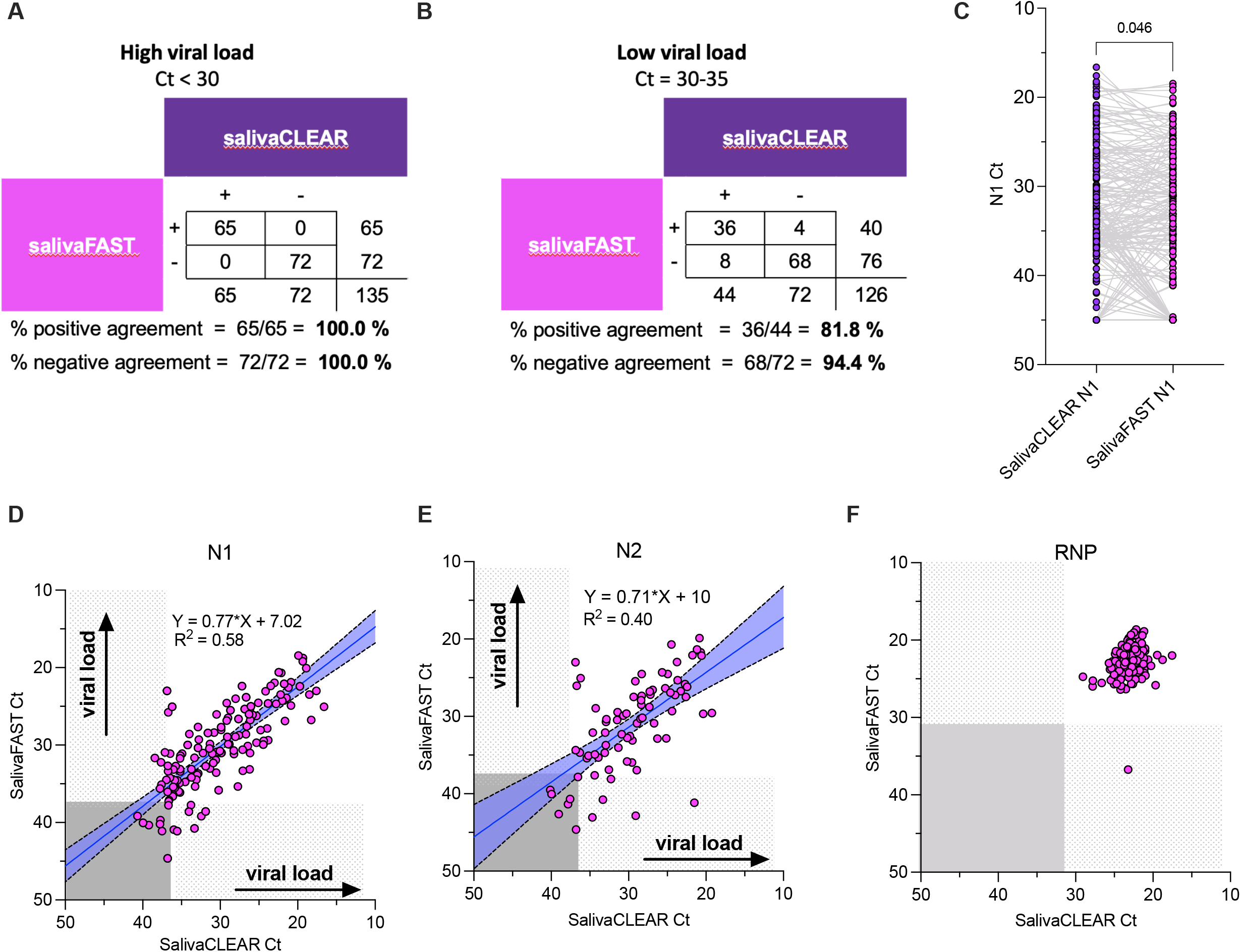
Impact of RNA extraction for RT-qPCR-based detection of SARS-CoV-2 in saliva. Positive and negative agreement between RNA extract from saliva (SalivaCLEAR) and extraction-free saliva (SalivaFAST) was assessed for high viral load nasal swab samples with a Ct<30 (A) and low viral load nasal swab samples Ct = 30-35 (B). Comparison of N1 Ct values from matched SalivaCLEAR and SalivaFAST samples demonstrated slightly lower viral load by SalivaFAST compared to SalivaCLEAR (C). There was moderate correlation between N1 Ct values (D) and N2 Ct values (E) for SalivaCLEAR and SalivaFAST. RNP values are relatively consistent for SalivaCLEAR and SalivaFAST (F).

### Comparison of primer/probe sets

The 2019-nCoV CDC assay for detection of SARS-CoV-2 implements two primer/probe pairs targeting the nucleocapsid gene (N1 and N2). Comparison of the performance of N1 and N2 within a single assay revealed a high degree of linearity for SwabCLEAR (*R*^*2*^=0.97), SalivaCLEAR (*R*^*2*^=0.83), and SalivaFAST (*R*^*2*^=0.82) (Fig. 5A-C), suggesting that most COVID-19 cases were detected by both targets with similar assessment of viral load. Whereas N1 detected several low viral load COVID-19 cases missed by N2, especially in saliva, N2 offered no additional diagnostic value.

**Figure 5.**
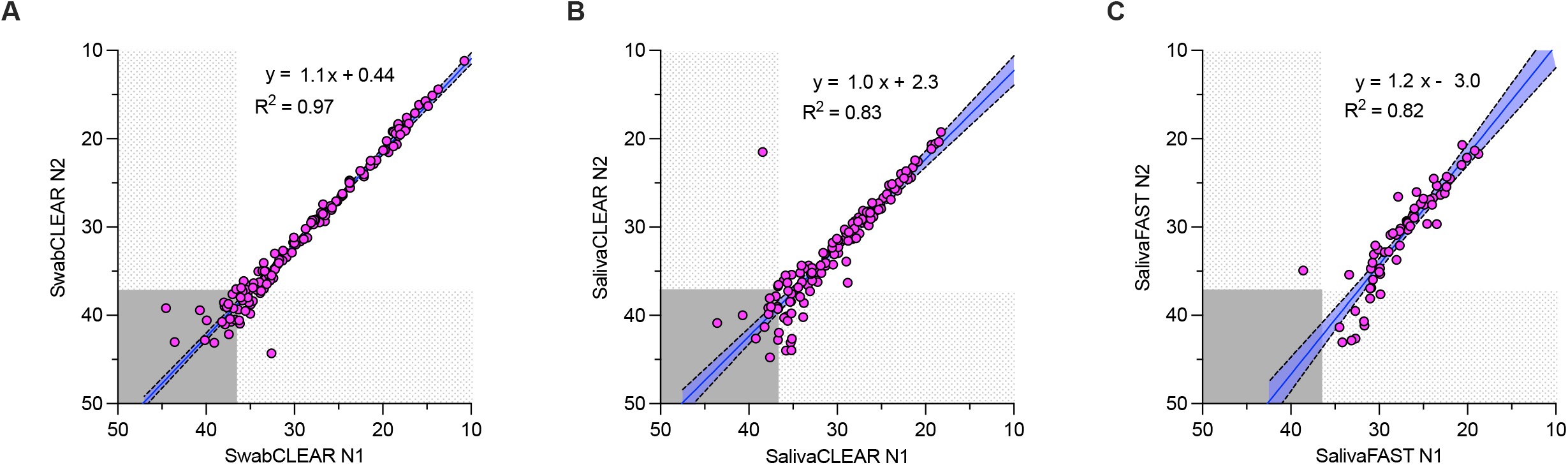
Comparison of N1 and N2. There is excellent correlation between Ct values for N1 and N2 for SwabCLEAR (A), SalivaCLEAR (B), and SalivaFAST (C) and negligible additional clinical sensitivity added by N2.

### Quantitation of viral load by RT-qPCR and ddPCR

To evaluate the quantitative accuracy of viral load by RT-qPCR, ddPCR using the same primers and probes for N1 and N2 was also performed on RNA extracts from nasal swab and saliva specimens. First, a standard curve was generated for SalivaFAST

First, quantitative accuracy and precision by ddPCR was performed. The recovered viral load in spiked samples following RNA extraction was 36.3-47.0% for nasal swab and 28.9-45.7% for saliva. Precision for N1 and N2 by ddPCR of extracts from nasal swab and saliva demonstrated a coefficient of variation (CV) of 5.6-12.3% and 4.5-19.1%, respectively (Fig. 1A). Compare of viral loads in specimen pairs according to N1 and N2 generally demonstrated no correlation (Fig. 6B-C). In nasal swab specimens, N1 demonstrated a broad range of viral load whereas N2 in the same samples plateaued (Fig. 6D-E), again demonstrating inferior performance of N2. This pattern was not observed in saliva which was likely related to overall lower viral loads (Fig. 6G-H, *R*^*2*^=0.87).

**Figure 6.**
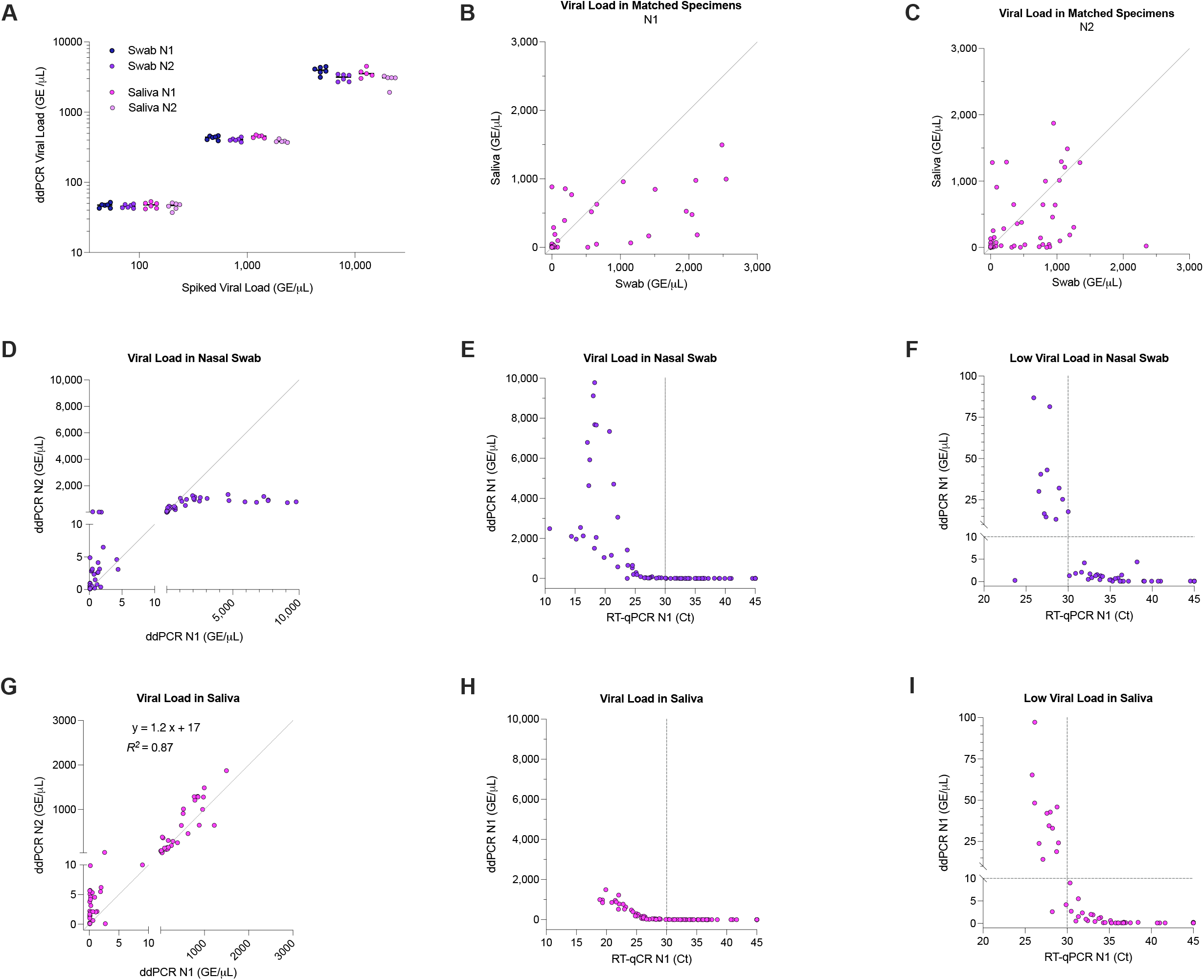
Evaluation of viral load according to RT-qPCR and ddPCR. There is excellent precision and systematic reduction in accuracy in viral load assessment by ddPCR of RNA extracts from both nasal swab and saliva samples (A). The viral loads of matched nasal swab and saliva specimens are compared according to N1 (B) and N2 (C). The performance of N1 and N2 is compared in individual nasal swab (D) and saliva (G). The N1 signal in RT-qPCR and ddPCR demonstrated a strong linear relationship for Ct values less than 30 (Fig. 6E, H). At low SARS-CoV-2 concentrations (<100 GE/µL) Ct values in nasal swab (Fig. 6F) and saliva (Fig. 6I) do not correlate with viral load.

Comparison of Ct values by RT-qPCR with viral load by ddPCR revealed a logarithmic pattern with a clear inflection point for both nasal swab specimens (Fig. 6E,H). Ct values below this inflection point demonstrated linear agreement in both assays with better correlation for saliva compared to nasal swab specimens. Ct values above this inflection point corresponded to low viral load (<10 GE/µL) (Fig. F,I). Taken together, these data confirm that viral loads are higher in nasal swab samples and that Ct values no longer have quantitative value when greater than 30.

### A combined nasal swab and saliva specimen is a viable specimen type

Given that RNA extraction is expendable for SARS-CoV-2 testing of saliva (Fig. 4A) and that viral load is slightly higher in nasal swab specimens compared to matched saliva specimens (Fig. 1A, Fig. 3C-E, Fig. 6), we hypothesized that extraction-free testing of combined nasal swab and saliva specimens (i.e., Spit-N-Swab) would be an acceptable approach for detection of SARS-CoV-2. COVID-19 patients provided saliva specimens in which nasal swabs were immersed. These specimens were tested directly without RNA extraction. Spit-N-Swab demonstrated a Ct increase of 1.6 compared to SwabCLEAR (n=104 specimen pairs, *p* = 0.011) and a Ct decrease of 1.1 compared to SalivaFAST (n=90 specimen pairs, Fig. 3A, *p* < 0.001) (Fig. 7A). Spit-N-Swab may improve the clinical sensitivity of SalivaFAST, given that low viral load was detected in some Spit-N-Swab specimens with a negative matched SalivaFAST specimen (Fig. 7C). These data suggest that a combined nasal swab and saliva specimen is viable for extraction-free testing and may enhance sensitivity in COVID-19 patients with low viral load infections.

**Figure 7.**
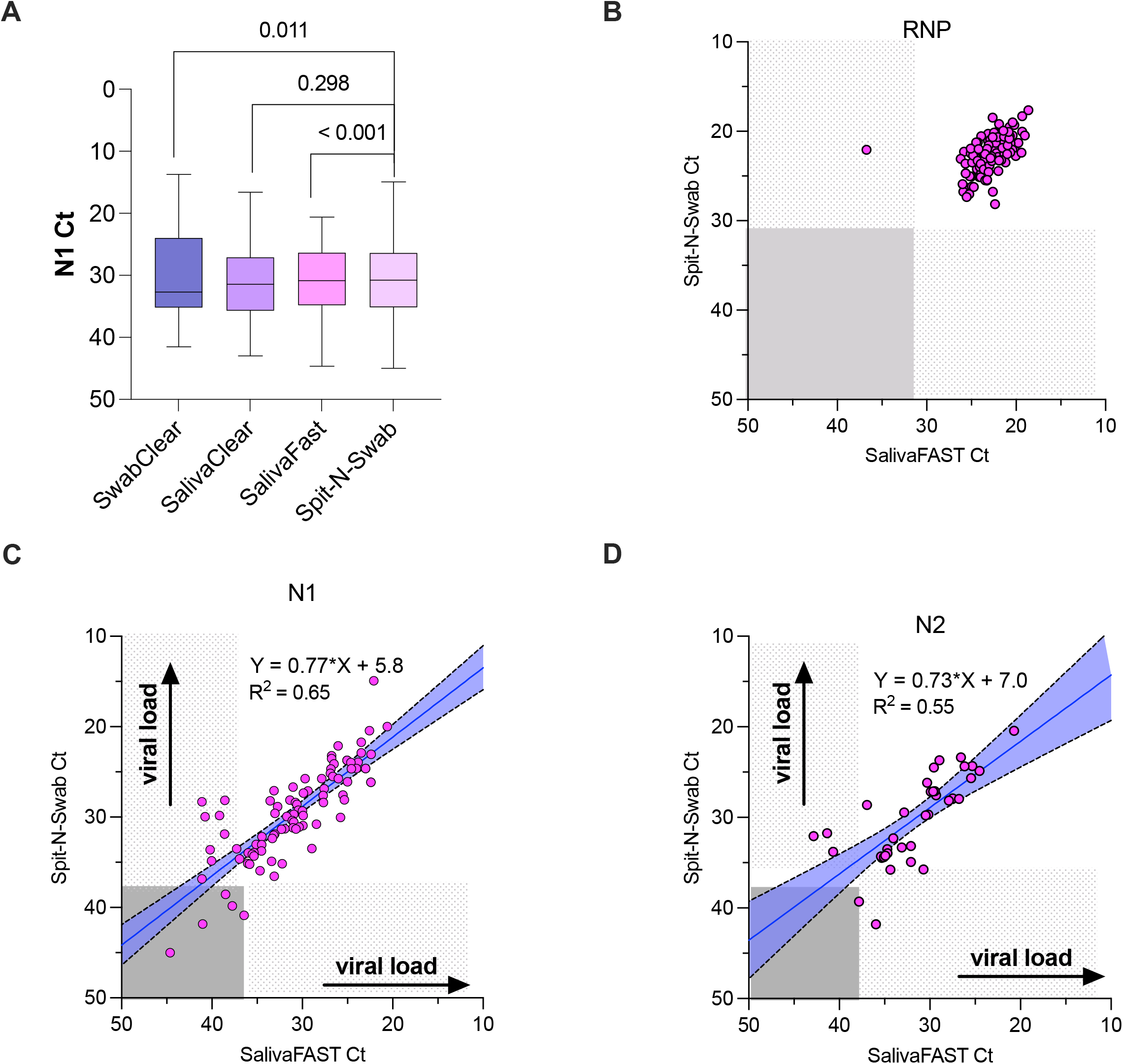
Extraction-free testing of saliva and combined nasal swab/saliva specimens. The N1 Ct values are compared across SwabCLEAR, SalivaCLEAR, SalivaFAST, and Spit-N-Swab (combined collection of nasal swab and saliva) (A). There was moderate correlation between SalivaFAST and Spit-N-Swab testing according to N1 (C) and the N2 (D) Ct values. The N1 Ct value of Spit-N-Swab specimens demonstrated low viral load samples missed by SalivaFAST (C). RNP Ct values were consistent in both assays (B).

### Morning versus afternoon collection

COVID-19 patients underwent longitudinal monitoring up to 27 days after symptom onset to observe the natural course of convalescence and viral clearance (Fig. S1). Many matched sample sets were collected in the morning and afternoon on the same day. Comparison of the RNP Ct value in the morning versus the afternoon demonstrated a slight Ct value increase of 0.69 in SwabCLEAR and 1.1 in SalivaCLEAR (Fig. 8A-B), suggesting diurnal change in the sample matrix. However, this difference was not observed for SalivaFAST (Fig. 8C). There was a trend for afternoon specimens to have decreased viral load compared to morning saliva specimens. This change was most pronounced for SalivaFAST, which demonstrated an Ct increase of 2.78 for N1 (*p* = 0.011 and 2.58 for N2 (*p* = 0.031)(Fig. 8C). Overall, these preliminary data raise the possibility that diurnal variation in saliva matrix may could impact the clinical sensitivity of RT-qPCR.

**Figure 8.**
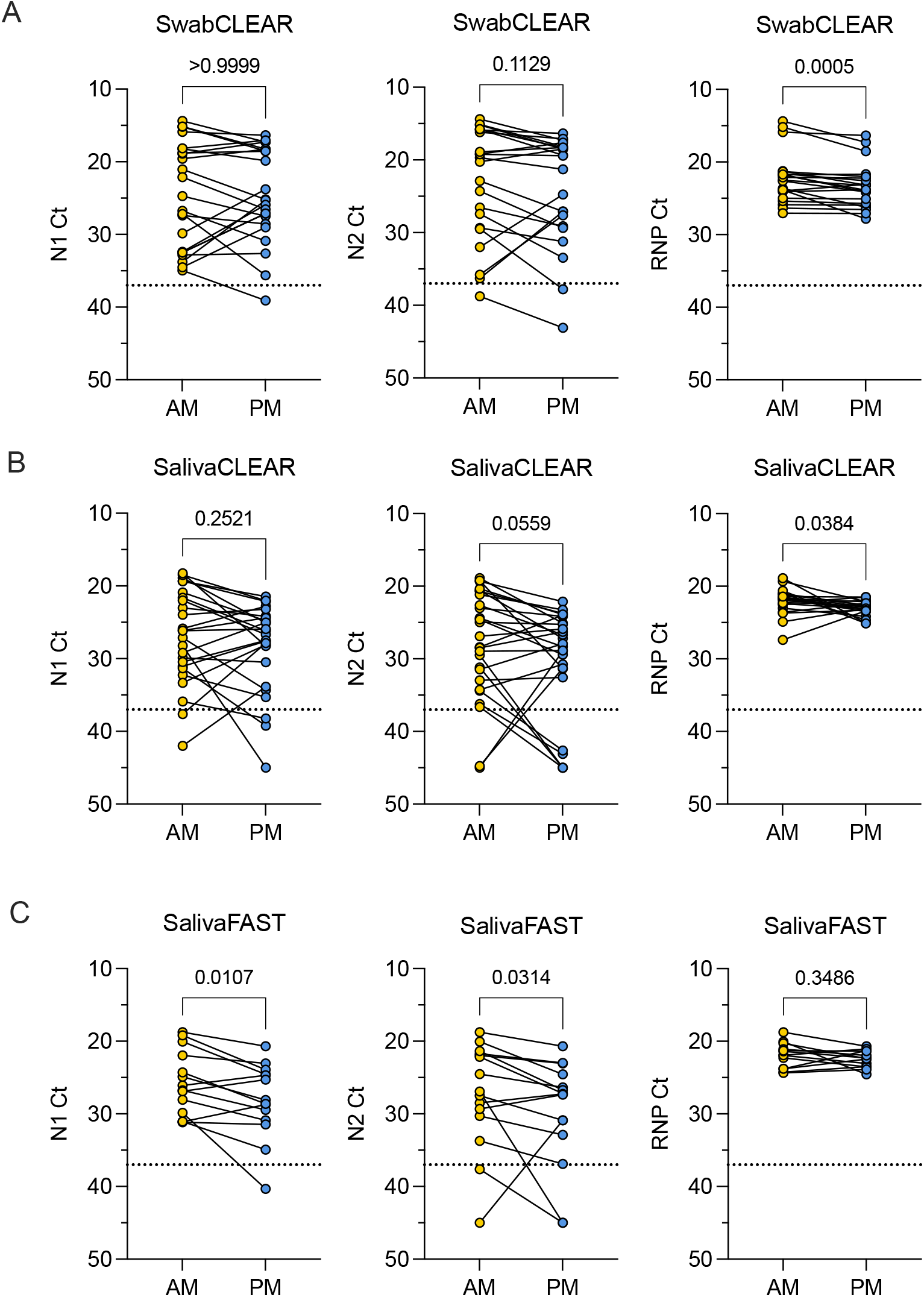
Impact of morning versus afternoon collection. Morning (AM, yellow) and afternoon (PM, blue) samples are compared for SwabCLEAR (A), SalivaCLEAR (B), and SalivaFAST (C). RNP Ct values were slightly lower in the PM compared to the AM for SwabCLEAR (A, right panel) and SalivaCLEAR (B, right panel). SalivaFAST demonstrated a similar trend that was not significant (C, right panel). Whereas the N1 Ct values did not differ significantly between AM and PM for SwabCLEAR (A, left panel) and SalivaCLEAR (B, left panel), SalivaFAST demonstrated significantly increased N1 (C, left panel) and N2 (C, center panel) Ct values in the PM compared to the AM. Assay threshold for N1 and N2 targets are represented by horizontal dotted lines.

## Discussion

We performed viral load monitoring by RT-qPCR testing of 231 matched specimen sets from COVID-19 patients in a prospective longitudinal study. Four specimen types were assessed: RNA extract from nasal swab (SwabCLEAR), RNA extract from saliva (SalivaCLEAR), saliva without extraction (SalivaFAST), and combined saliva and nasal swab without extraction (Spit-N-Swab).

Nasal swab and saliva are comparable specimens for detection of SARS-CoV-2. Although viral load was generally higher in nasal swab compared to matched saliva, this difference diminished over the course of infection and had no impact on clinical sensitivity (Figs. 1, 3, 6). Extraction-free testing of combined nasal swab and saliva resulted in slightly lower Ct values compared to extraction-free saliva testing (Fig. 7), further supporting higher viral load in the nasal cavity compared to the oral cavity. Although RNA extract from saliva demonstrated a slightly increased viral load compared to extraction-free saliva, there was excellent positive agreement between matched specimens (Fig. 4). These findings suggest that RNA extraction is an expendable step for RT-qPCR testing for SARS-CoV-2, as others have observed (*8, 17*). Extraction-free RT-qPCR represents a new clinical testing paradigm during the COVID-19 pandemic. Saliva is an emerging specimen type that could also have utility for other respiratory pathogens.

Quantitation of viral load has yet to be realized for COVID-19 testing. Increased viral load has been reported in nasopharyngeal (NP) swabs compared to saliva, with conflicting data on overall sensitivity (*17-28*). Others have observed higher viral load in saliva and even improved clinical sensitivity compared to NP swab (*8, 29*). These inconsistencies may reflect pre-analytical variables that introduce artificial differences, such as collection device, or reveal the dynamics of SARS-CoV-2 infection in the nasal and oral cavities. Most relevant professional groups including American Public Health Labs, Infectious Disease Society of America, Association of Molecular Pathology, College of American Pathology, FDA and CDC currently oppose reporting of Ct values as an indicator of viral load (*1*). Indeed, the FDA has authorized laboratories performing SARS-CoV-2 RT-qPCR assays with EUA to provide qualitative interpretation as positive or negative, although laboratories can choose to report Ct values (*2*). We observe that RT-qPCR approximates the viral load in saliva such that a Ct value of 20 corresponds to a viral load of 1,000 GE/µL, and Ct value of 26 corresponds to a viral load of 100 GE/µL, and Ct values >30 represent viral loads near the assay LoD at 4 GE/µL. The same general observations were observed for nasal swab samples, with much less accuracy at high viral loads (Fig. 6). These data suggest that viral load monitoring of an individual patient, using the same specimen type and RT-qPCR assay and performed by the same laboratory, may have value, for example to personalize quarantine schedules or guide care of a hospitalized patient.

Some indirect evidence suggests that saliva may harbor virus late in infection or shed non-infectious viral particles following viral clearance in the nasal cavity. For example, a meta-analysis of matched saliva and NP swab studies demonstrated that the sensitivity of saliva was higher than NP swab in known COVID-19 patients, but lower in individuals not previously diagnosed with COVID-19 (*24*). We did not observe this trend (Fig. S1). Moreover, saliva is not simply a surrogate for sampling of the nasal cavity. Epithelial cells in salivary glands and oral mucosa express viral entry factors that permit SARS-CoV-2 infection, and these infected cells can be detected in saliva (*30, 31*). The timing of saliva collection may also affect SARS-CoV-2 testing, as we observed slightly higher viral load in saliva collected in the morning compared to the afternoon (Fig. 8C). This may be related to exogenous inhibitory substances consumed throughout the day or diurnal variation in saliva secretion. Overall SARS-CoV-2 infects the oral cavity and saliva represents a unique window into SARS-CoV-2 infection.

This prospective longitudinal study design observed COVID-19 patients through convalescence and resulted in an abundance of low viral load specimens, which are particularly useful to stress RT-qPCR assays near the clinical decision threshold. However, there are limitations of this study. First, this study did not include collection of matched NP swabs and compared the relative performance of nasal swabs and saliva. Second, the small cohort of community COVID-19 patients studied here is likely not representative of other patient populations, such as immunocompromised or hospitalized patients. Third, this study was performed in 2020 prior to the emergence of SARS-CoV-2 variants of concern in the U.S. and widespread COVID-19 vaccination. At that time the alpha variant predominated in the studied geographic region. The delta variant for example may display altered biology. Yet RT-qPCR remains foundational for SARS-CoV-2 testing. Simplified specimen collection using saliva and efficient laboratory testing using extraction-free methods offer an innovative approach that is attractive to resource-poor settings and high throughput laboratories alike. Viral load monitoring of individual patients by a single laboratory has the potential to predict outcomes in COVID-19, assess therapeutic response particularly in the setting of clinical trials, and personalize guidance for quarantine and repeat testing.

## Materials and Methods

### Patient recruitment and sample collection

Patients with COVID-19 symptoms, specifically for the alpha variant, or known close contacts of an infected individual were recruited to provide nasal swab and saliva specimens as part of an IRB-approved study. COVID-19 status was confirmed by an alternative method with FDA EUA. Nasal swab collection of anterior nares for SwabCLEAR testing was performed using the DNAGenotek ORE-100 device (*32*). Saliva collection was performed using either the DNAGenotek OM-505 device (*33*) for SalivaCLEAR or Falcon 50 mL conical tubes or cryotubes outfitted with a saliva collection aid (Salimetrics) for SalivaFAST. Patients also provided a combined nasal swab and saliva specimen for Spit-N-Swab in which nasal swab was placed into saliva. Patients underwent longitudinal sample collection until viral clearance and/or withdrawal from the study (Fig. S1). All samples were transported to the lab at ambient temperature within 72 hours and analyzed on the day of receipt in the laboratory.

### RT-qPCR for detection of SARS-CoV-2

The CDC 2019 Novel Coronavirus (2019-nCoV) Real-Time RT-qPCR assay with primers and probes from Integrated DNA Technologies (IDT) was implemented. Briefly, this assay includes two primer/probes (N1 and N2) for detected of SARS-CoV-2 and one primer/probe for detection of ribonuclease P (RNP) (*15*). RNA extraction of SwabCLEAR and SalivaCLEAR samples was performed using QIAamp Viral RNA mini Kit (Qiagen). Saliva samples collected in 50 mL falcon tubes or in a cryotube were tested directly by extraction-free SalivaFAST testing. 5 µL of swab RNA extract, 5 µL of saliva RNA extract, or 5 µL of raw saliva was added to PCR master mix to a volume of 20 µL into 96-well plates (BioRad). RT-qPCR was performed by Bio-Rad CFX Connect Real-Time PCR Detection System with the CFX software. Cycle conditions were 55°C for 10mins; 95°C for 1min; and 95°C, 10s; 60°C, 30s for 45 cycles. For analytical validation and assay controls, synthetic SARS-CoV-2 nucleic acid was used (Twist Biosciences).

### ddPCR for detection of SARS-CoV-2

Viral load quantitation by the Bio-Rad SARS-CoV-2 digital droplet PCR (ddPCR) kit containing the 2019-nCoV CDC ddPCR triplex probe assay was performed on a Bio-Rad Automated Droplet Generator and QX200 Droplet Reader according to manufacturer instructions (*34*). The reported LoD of this assay is 0.150 copies/µL.

### Data Analysis

Data was analyzed using Microsoft Excel and GraphPad Prism Version 9.1.1. A two-tailed Wilcoxon signed rank test was applied for matched specimen analysis and a Mann Whitney test was used to compare unpaired groups.

## Data Availability

Data is presented in the manuscript

## Acknowledgments

The authors would like to thank CU Innovations and the Stem Cell Biobank and Disease Modeling Core at the University of Colorado Anschutz Medical Campus.

## Author contributions

Conceptualization (YQ, BB, XY, JZ, BH, SL)

Investigation (YQ, LL, PM, CH, FG, AH, RT, SY, XY, BH, SL)

Methodology (YQ, LL, PM, CH, IK, BH, SL)

Data Curation (YQ, LL)

Formal Analysis (DG, PM, BH)

Writing – original draft (BH)

Writing – review & editing (XY, SL, BH)

Resources (SL)

Funding acquisition (SL)

## Data and materials availability

A master transfer agreement (MTA) was not used to access samples in this study. All data associated with this study are available in the main text or the supplementary materials.

## Figure Legends

**Figure S1.**
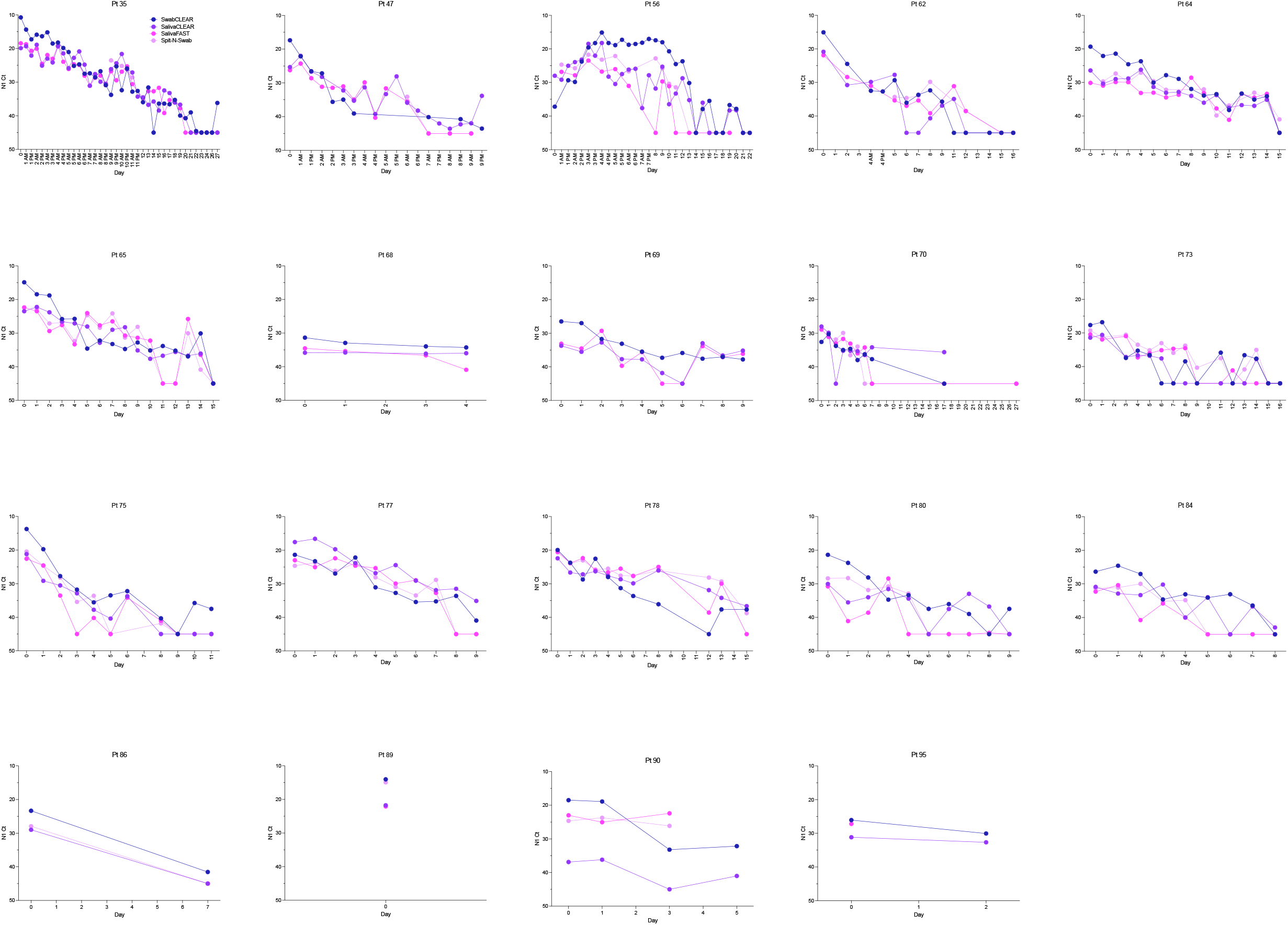
Viral load monitoring in COVID-19 patients from diagnosis to viral clearance. Ct values are presented for matched SwabCLEAR (blue), SalivaCLEAR (purple), SalivaFAST (magenta), and Spit-N-Swab (pink) specimens collected longitudinally over 5 to 21 days from COVID-19 patients.

